# Limited Generalizability of Epigenetic Clocks and Scores Across Pediatric Tissues and Age Ranges

**DOI:** 10.64898/2025.12.11.25340872

**Authors:** Vera N. Karlbauer, Carmen Domínguez-Baleón, Monika Rex-Haffner, Tamara Namendorf, Ferdinand Hoffmann, Heiko Klawitter, Sonja Entringer, Claudia Buss, Sibylle M. Winter, Christine Heim, Elisabeth B. Binder, Darina Czamara

## Abstract

Epigenetic clocks and scores have been investigated as potential biomarkers of later-life health outcomes following early-life exposures. In pediatric settings, DNA methylation (DNAm) is often measured in saliva; however, most clocks and scores have been trained in adult blood. Therefore, we assessed the performance, correlation, longitudinal stability, and association with early-life adversity (ELA) for established epigenetic measures in matched pediatric blood and saliva samples. Leveraging the Kids2Health cohort of 291 children (3-12 years, 56% exposed to ELA), we assessed DNAm (Illumina EPICv2) from matched blood and saliva. We calculated 22 commonly used epigenetic measures (chronological and biological clocks, scores for body mass index [BMI], C-reactive protein [CRP], cognition, maternal smoking, telomere length) and compared them with corresponding measured phenotypes (chronological age, BMI, CRP, IQ, telomere length). Overall, performance of epigenetic clocks and scores in children varied widely. Nine epigenetic measures were significantly and equally correlated with their corresponding phenotype across tissues. Six measures were highly correlated between blood and saliva (r≥0.7). All age acceleration estimates showed low to moderate cross-tissue correlations (r=0.20–0.68). Epigenetic scores indicating lower cognitive ability and elevated inflammation were associated with ELA and low SES in both tissues. We additionally provide epigenome-wide blood-saliva correlations across 815,069 CpGs. The results indicate limited generalizability of adult-trained epigenetic clocks and scores to pediatric blood and saliva, even when accounting for cell type composition. We advise caution for cross-tissue and cross-age-range applications of epigenetic measures in research and clinical settings and provide a resource to optimize epigenetic biomarkers in children.

## Introduction

DNA methylation (DNAm) is frequently investigated as a pathway linking environmental exposures to later-life health outcomes ^1^. Epigenetic clocks use DNAm to estimate biological age, which can deviate from chronological age ^2^, and accelerated epigenetic aging has been associated with diverse environmental exposures and increased risk for mental as well as physical disorders ^3^, while epigenetic age deceleration has been linked to positive health behaviors such as physical exercise and healthy nutrition ^4^. Beyond clocks, DNAm-based measures have been trained to predict exposures and phenotypes, e.g. smoking, body mass index (BMI), immune measures or cognitive ability, resulting in epigenetic scores (also called methylation profile scores or polyepigenetic scores), which have found increasing use in research ^5,6^.

Since adverse exposures occurring early in life, such as early-life adversity (ELA), socioeconomic disadvantage, or exposure to toxins, are associated with increased long-term disease risk ^7,8^, pediatric cohorts are central to studying the epigenetic embedding of exposures ^9,10^. In addition, measuring epigenetic biomarkers of long-term risk in childhood would allow targeted early prevention measures. However, most studies in pediatric populations rely on epigenetic measures developed in adult blood using linear models, which are then applied to pediatric blood or saliva samples, despite evidence of non-linear DNAm dynamics during childhood and adolescence ^11,12^. Applying linear adult-trained measures implicitly assumes that processes underlying mid- and late-life biological aging operate similarly during childhood and adolescence. Moreover, it is unclear whether epigenetic age acceleration or deceleration is linked to more favorable health outcomes in children ^9^. Furthermore, it remains to be evaluated whether epigenetic scores developed in adults, e.g. predicting BMI or C-reactive protein (CRP), also translate to children, where these measures might have different epigenetic correlates in the context of development.

The use of epigenetic measures in children is further complicated by often mismatched training and target tissues: Many clocks and scores have been trained in blood, whereas pediatric studies often collect saliva due to ease of sampling and storage. According to a recent meta-analysis on epigenetic aging and social determinants of health, 35% of pediatric epigenetic studies used saliva compared with 16% in adults ^13^. However, DNAm is highly tissue- and cell-type-specific ^14^, a problem long recognized in epigenetic research as the ‘tissue issue’ ^15^. Not all tissues might age or respond to exposures and diseases similarly ^16;17^. These concerns are amplified in children due to differences in cell type composition. Since DNAm is typically measured in bulk tissue, cell type heterogeneity drives variation in DNAm data and affects epigenetic clock and score estimates ^18^. Blood primarily contains immune cells, whereas saliva is a mixture of immune and buccal epithelial cells (BECs). In children, immune cell proportions in saliva are lower than in adults and increase with age ^19,20^. Immune cells and BECs differ in their DNAm profiles ^21^, and saliva with a higher immune cell content more closely resembles blood ^22^, making salivary and blood DNAm less concordant in children than in adults. While cell type composition of blood and saliva samples can be estimated from DNAm data ^23^ and is usually controlled for statistically, it is unclear whether this adjustment sufficiently reduces tissue differences ^24,25^.

These issues can be mitigated by using age- and tissue-matched epigenetic measures, such as PedBE ^26^ (BECs, 0–20 years) or Wu’s blood-based clock ^27^ (0–18 years), but their use in research remains limited. In fact, the aforementioned meta-analysis reported that 60% of pediatric studies used epigenetic clocks trained exclusively in adult samples ^13^. In addition, current pediatric clocks that are accurate predictors of chronological age might not be sensitive to exposures or health-relevant phenotypes, as a recent study showed that PedBE was neither associated with socioeconomic status (SES) or health outcomes in young adults ^28^. Consequently, many pediatric studies rely on measures that lack systematic evaluation of reliability and generalizability in children.

The use of epigenetic measures in tissues and age groups that were not part of their original training sample makes it difficult to be confident about their clinical utility. To inform health interventions and clinical practice, clocks and scores need to first be validated in the respective age group and tissue. For example, GrimAge, an epigenetic clock trained in adults of 40 or older ^29^, was proposed for the identification of children at risk for obesity after surviving cancer ^30^ and the DunedinPACE clock, trained in 26-45-year-old adults ^31^, recommended for monitoring cardiometabolic health in children and adolescents ^32^. Without prior validation, it is unclear whether these associations should inform health interventions. Similarly, applying adult blood-trained measures to youth saliva to evaluate the effectiveness of health interventions, as previously proposed ^33,34^, might bias assessments with implications for policies affecting vulnerable populations such as socioeconomically disadvantaged youths.

Epigenetic measures are a promising tool for identifying biological signatures of ELA years before the onset of disease. But before drawing far-reaching conclusions about their use in clinical care and intervention, we need to first evaluate their generalizability across ages and tissues. So far, neither performance nor cross-tissue correlations of epigenetic measures have been systematically assessed in large pediatric populations with matched cross-tissue samples. Two recent studies ^24,25^ investigated blood-saliva correlations for several epigenetic clocks and scores, but their sample sizes were limited and included few children and adolescents (5–36 samples from children under 18 years).

To address this issue, we leverage a cohort of 291 children aged 3-12 years with matching blood and saliva samples to (1) evaluate the performance of established epigenetic measures in pediatric samples via association with measured phenotypes (2) explore cross-tissue correlations and longitudinal stability of epigenetic measures, (3) compare the associations of ELA and SES with these epigenetic measures in blood and saliva, and (4) provide an epigenome-wide database of correlations between pediatric blood and salivary DNAm as a resource for the development of future epigenetic measures.

## Methods

### Study population

We used a subsample of 291 children aged 3–12 years from the Kids2Health cohort, a Berlin-based case-control study for studying the mechanisms of biological embedding of ELA. The full cohort included 245 children with exposure to childhood maltreatment, 49 children with war-and migration-related ELA, and 241 controls without exposure. The sample was enriched for clinically obese children. 291 children were selected for analysis based on availability of matching blood and salivary DNAm. We additionally included longitudinal DNAm data from a one-year follow-up (mean lag=1.23 years) for 70 children with blood DNAm (52 of which were part of the cross-tissue baseline sample) and 85 children with saliva DNAm (73 of which were part of the cross-tissue baseline sample). Ethical approval for the study was granted by the Ludwig-Maximilians-Universität München (LMU) Ethics Committee (approval # 18-444). All procedures were in accordance with the Ethical Principles for Medical Research established by the Declaration of Helsinki ^35^. Written informed consent was obtained from the children directly or from caregivers if children were younger than six years. Further cohort and recruitment details can be found in *eMethods*.

### Measures

#### DNA methylation

Peripheral blood and saliva were sampled in the morning of the same measurement day, except for three individuals with a maximum difference of 35 days between samples. Blood and salivary DNA were extracted, run on the EPIC v2.0 Bead Chip (Illumina Inc., San Diego, USA), and pre-processed separately for each tissue. After quality control, 291 common baseline samples and 815,069 common CpGs across tissues remained. See *eMethods* for further details.

#### Epigenetic clocks and scores

We calculated a set of epigenetic clocks frequently used in research, including first-generation chronological clocks trained to predict chronological age (Horvath multi-tissue ^36^, HorvathSkinBlood ^37^, Hannum ^38^, PedBe ^26^, and Wu’s clock ^27^), second-generation biological clocks trained to predict health-relevant biomarkers (PhenoAge ^39^, GrimAge ^29^, and GrimAge2 ^40^), and third-generation biological clocks trained to predict the pace of biological aging (DunedinPACE ^31^). We also calculated principal component-based versions (PC clocks) of the Horvath multi-tissue, HorvathSkinBlood, Hannum, PhenoAge, and GrimAge clocks that were re-trained on principal components of multiple CpGs for increased stability ^41^. For all clocks except DunedinPACE, we obtained two measures of epigenetic age acceleration: *a)* the residual of estimated epigenetic age against chronological age (accel) and *b)* the difference of estimated epigenetic age minus chronological age (delta). Since residuals are preferable to deltas in most research scenarios ^42^, we will only discuss acceleration residuals. Additionally, we computed epigenetic scores by summing the product of CpG weights derived from epigenome-wide association studies (EWAS) for the phenotype of interest with individual DNAm values. We calculated two epigenetic scores for BMI ^43,44^, two epigenetic scores for CRP ^45,46^ and an epigenetic score for cognitive ability (epigenetic-g) ^47^. We also computed epigenetically estimated leukocyte telomere length (DNAmTL) ^48^, its PC-based version ^41^, and an epigenetic score for prenatal smoking exposure ^49^. All measures were estimated separately per tissue. We used tissue-specific reference panels ^21,50^ for cell type deconvolution. Measures were residualized for estimated cell type proportions, and analyses were performed on unresidualized and cell-type-residualized measures. For additional information on clock/score characteristics, selection and calculation, see *eTable 1* and *eMethods*.

#### Salivary and blood biomarkers

CRP levels (N=196) were determined from saliva and telomere length (T/S ratio, N=286) was determined from blood DNA via qPCR (see *eMethods)*.

#### Early-life adversity and socioeconomic status

Exposure to ELA was coded as a dichotomous variable (exposed vs. unexposed). Children were classified as ELA-exposed if they experienced childhood maltreatment as determined by the Maternal Interview for the Classification of Maltreatment (MICM) ^51^ or war- and migration-related traumatic events according to the UCLA post-traumatic stress disorder reaction index ^52^. Family SES was determined from parental education, vocation, and family income ^53^. See *eMethods* and previous publications on the cohort ^54,55^ for further information.

### Statistical analysis

All statistical analyses were performed in R version 4.5.0 ^56^ and RStudio ^57^. Analyses were carried out for uncorrected as well as cell-type-corrected epigenetic measures and p-values were FDR-corrected to account for multiple testing. Due to the presence of related pairs in the sample, all regression models containing between-person effects were adjusted for genetic kinship based on genotype data. See *eMethods* for details.

In order to examine clock performance in blood and saliva, we first calculated the median absolute error (MAE) for each clock reflecting the deviation between estimated epigenetic and chronological age in years. Additionally, we tested whether the performance of epigenetic clocks and scores, i.e. the association between an epigenetic measure and its corresponding phenotype, differed between blood and saliva. We ran linear models predicting each epigenetic measure from the corresponding phenotype, tissue and a phenotype-by-tissue interaction term. A significant phenotype-by-tissue interaction indicates tissue differences in the association of epigenetic measure and corresponding phenotype. We tested estimated epigenetic age on all clocks except for DunedinPACE and used chronological age as corresponding phenotype (N=291). For epigenetic BMI, we measured BMI (N=290). Salivary CRP levels (N=196) were used as phenotype for epigenetic CRP, and blood telomere T/S ratio (N=286) was used as phenotype for epigenetic telomere length. For epigenetic-g, nonverbal IQ was chosen (N=287).

For assessment of cross-tissue comparability, we calculated within-person Pearson correlations between blood and saliva for epigenetic aging measures (age, accel, and delta), and epigenetic scores for all blood-saliva pairs at baseline. Next, we tested the longitudinal stability of epigenetic measures via within-person Pearson correlations between baseline and one-year follow-up in blood (N=70) and saliva (N=85). We considered correlations of r<0.3 as low, r=0.3–0.7 as moderate, and r≥0.7 as high.

We then tested for cross-tissue consistency of ELA and SES associations with epigenetic age acceleration and epigenetic scores. For each epigenetic measure in each tissue, we performed linear regressions with the epigenetic measure as outcome and ELA or SES as predictor. Models were adjusted for age, genetically determined sex, genetic kinship, an epigenetic proxy for cigarette smoke exposure via cg05575921 ^58^, and tissue-specific cell type proportions, and all predictors were standardized.

Moving beyond aggregated measures to epigenome-wide analyses, we calculated pairwise within-person Spearman correlations between blood and saliva for 815,069 CpGs that were available post-QC in both tissues. CpGs were deemed highly correlated if they were variable according to the definition of Hannon et al. (difference in DNAm between 10^th^ and 90^th^ percentile across all samples >5%) ^59^ and showed a cross-tissue correlation of ρ>.5. A set of follow-up analyses was performed to assess the function of highly correlated and variable CpGs (see *eMethods*).

## Results

### Sample characteristics

The cross-tissue baseline sample was aged 3–12 years (mean=8.12. SD=2.14), 45.02% female, and 56.01% exposed to ELA (see *eTable 2*). The most common self-reported ethnicity category was White (81.33%).

### Performance of epigenetic clocks in blood and saliva

We first compared the performance of epigenetic clocks (mostly trained in adult blood) in pediatric blood and saliva using absolute agreement with chronological age (MAE) and relative performance from linear models of tissue, age, and their interaction. As shown in *eFigure 1*, MAEs ranged widely from 0.95–44.42 years. Horvath’s SkinBlood clock achieved the best performance in blood (MAE=0.95 years), and PedBE in saliva (MAE=0.97 years).

Statistical comparison of age predictions between the tissues revealed two patterns exemplified in *Figure 1*. The biological clock GrimAge2, trained in adult blood, represents scenario 1. Without adjustment for cell type composition, the association between chronological and epigenetic age was significantly stronger in blood than saliva (*Figure 1A*; r=0.46 vs. 0.11, interaction p_fdr_<.001). However, after cell type correction, this interaction became non-significant, with a decreased correlation in blood and an improved correlation in saliva (*Figure 1B*; r_blood_=0.33, r_saliva_=0.43, interaction p_fdr_=0.43). This feature was shared by Horvath, SkinBlood, Wu’s clock, PhenoAge, and GrimAge (see *eFigure 2* and *eTable 3*). In summary, clocks in scenario 1 showed comparable performance in pediatric blood and saliva after adjustment for cell type composition.

Scenario 2 is represented by the PedBE clock, trained in children on salivary BECs. PedBE showed clear performance differences between blood and saliva (*Figure 1C*; r_blood_=0.44, r_saliva_=0.88, interaction p_fdr_<.001), which could not be attenuated by cell type correction (*Figure 1D*; r_blood_=0.35, r_saliva_=0.85, interaction p_fdr_<.001). This scenario was also seen in all PC clocks (PCHorvath, PCSkinBlood, PCHannum, PCPhenoAge, and PCGrimAge) as well the Hannum clock, which all showed significant performance differences with and without cell type correction (interaction p_FDR_<.05). Interestingly, for most blood-trained clocks in scenario 2, the performance advantage of blood was *reversed* after cell type correction, with saliva significantly outperforming blood. Thus, clocks in scenario 2 exhibited clear tissue differences despite adjustment.

**Figure 1.**
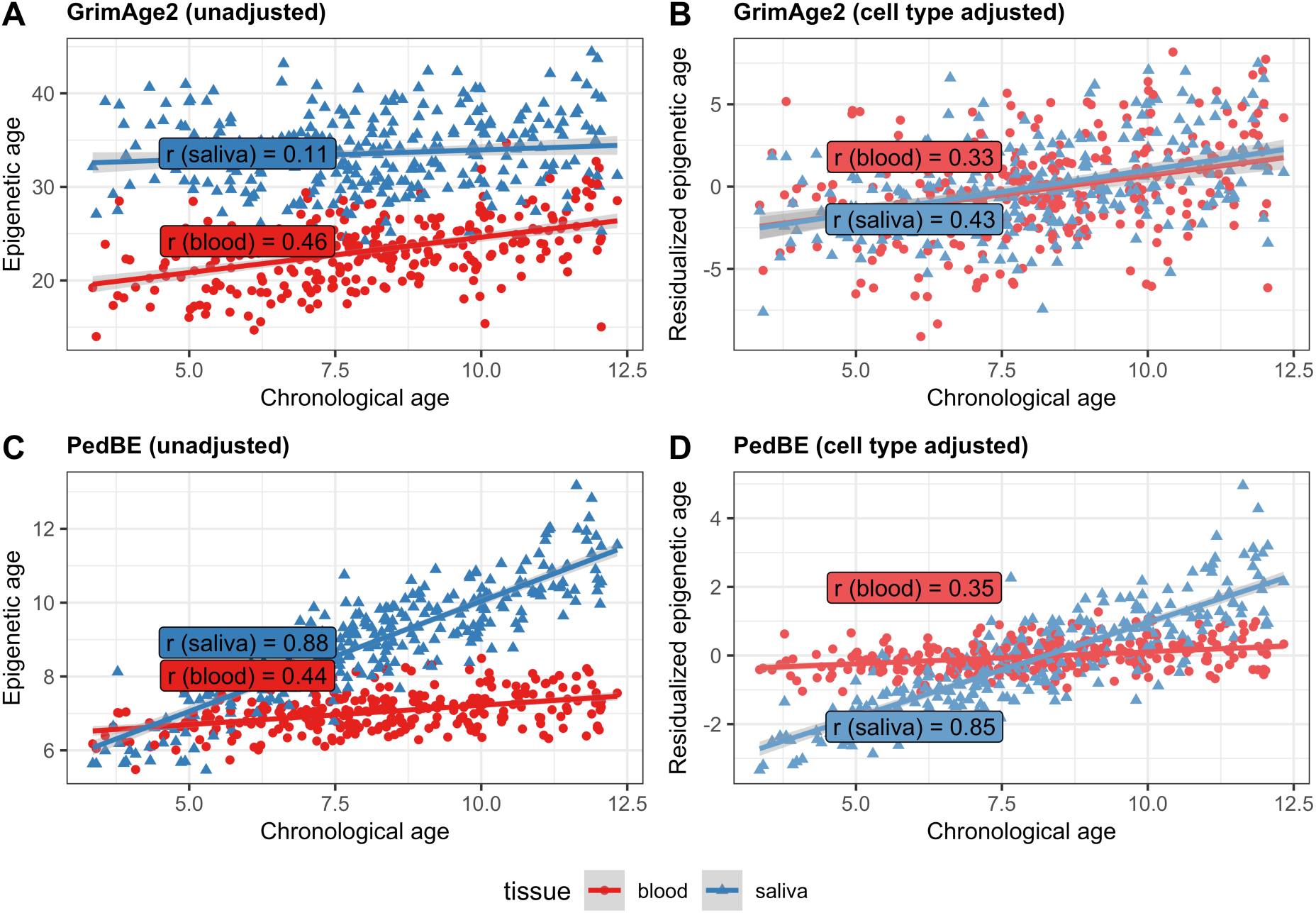
Performance of select epigenetic clocks by tissue. Association between chronological (x-axis) and epigenetic age (y-axis) by tissue for two select clocks, GrimAge2 (Panel A-B), trained on blood from adults aged 40-92, and PedBE (Panel C-D), trained on BECs from participants aged 0-20. Without cell type adjustment (Panel A), correlations between chronological age and GrimAge2 are significantly lower in saliva than in blood (r_blood_=0.46, r_saliva_=0.11, interaction p_fdr_<.001). After adjustment for estimated cell types (Panel B), associations with chronological age are comparable between blood and saliva (r_blood_=0.33, r_saliva_=0.43, interaction p_fdr_=0.43). For PedBE, saliva significantly outperforms blood (Panel C; r_blood_=0.44, r_saliva_=0.88, interaction p_fdr_<.001), and this tissue difference persists after cell type correction (Panel D; r_blood_=0.35, r_saliva_=0.85, interaction p_fdr_<.001). Similar plots and corresponding statistical results for all evaluated epigenetic clocks can be found in *e*Figure 2 and *eTable 3*.

### Performance of epigenetic scores in blood and saliva

We next evaluated seven epigenetic scores in pediatric blood and saliva: two BMI scores (Do et al. ^43^; Wahl et al. ^44^), two CRP scores (Wielscher et al ^46^; Ligthart et al. ^45^), epigenetig-g, and two scores trained on telomere length (DNAmTL, and PC DNAmTL). For these scores, target phenotypes were available in the Kids2Health cohort (BMI, salivary CRP, nonverbal IQ, leukocyte telomere length), see *eTable 4 and eFigure 3*. Correlations of epigenetic scores with their corresponding phenotypes were small to moderate (r=-0.02– 0.33, cell type corrected). When statistically comparing score performance between blood and saliva, phenotype-by-tissue interaction analyses also yielded two patterns. In scenario 1, epigenetic CRP (based on Wielscher et al.) was only associated with measured CRP in blood, but not in saliva prior to cell type correction (*Figure 2A*; r_blood_=0.26, r_saliva_=-0.04, interaction p_fdr_=.03). After adjustment, associations were similar across tissues, as reflected by a non-significant interaction (Figure 2B; r_blood_=0.19, r_saliva_=0.26, interaction p_fdr_=.05). Epigenetic BMI (based on Do et al.) and epigenetic-g showed the same pattern. In scenario 2, on the other hand, differences in score performance persisted after adjustment. DNAmTL was positively associated with measured telomere length in blood, but not in saliva (Figure 2C; r_blood_=0.14, r_saliva_=-0.13, interaction p_fdr_=.01). After cell type correction, both associations were non-significant (Figure 2D; r_blood_=0.06, r_saliva_=-0.02, interaction p_fdr_=.32). The same pattern applied to PC DNAmTL. Epigenetic BMI (based on Wahl et al.) and CRP (based on Ligthart et al.) also fell into scenario 2, as score performance differed significantly between blood and saliva with and without cell type correction. Similar to most epigenetic clocks in scenario 2, the pre-adjustment advantage of blood reversed post-adjustment (see *eTable 4* and *eFigure 3*).

**Figure 2.**
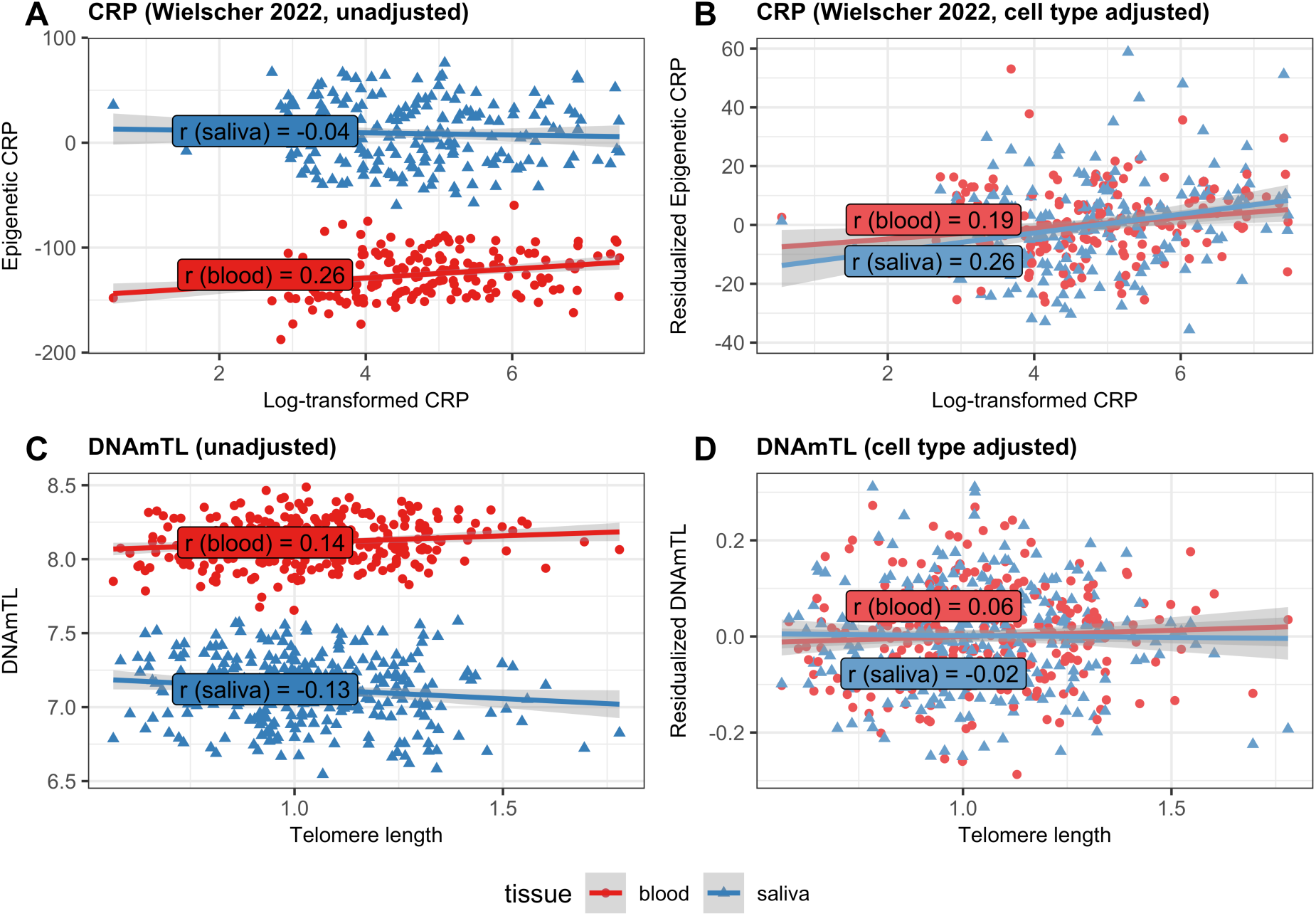
Performance of select epigenetic scores by tissue. Association between corresponding phenotype (x-axis) and epigenetic score (y-axis) by tissue for two select scores, CRP based on Wielscher (2022, Panel A-B), trained to predict salivary CRP from adult blood DNAm, and DNAmTL (Panel C-D), trained to predict adult leukocyte telomere length. Without cell type adjustment the correlation between measured and epigenetic CRP is only significant in blood (Panel A; r_blood_=0.26, r_saliva_=-0.04, interaction p_fdr_=.03). After adjustment for estimated cell types, associations are comparable between blood and saliva (Panel B; r_blood_=0.19, r_saliva_=0.26, interaction p_fdr_=.05). Unadjusted DNAmTL is only positively associated with telomere length in blood (Panel C; r_blood_=0.14, r_saliva_=-0.13, interaction p_fdr_=.01). After cell type correction, there is no significant association of DNAmTL and telomere length in either blood or saliva (Panel D; r_blood_=0.06, r_saliva_=-0.02, interaction p_fdr_=.32). Similar plots and corresponding statistical results for all epigenetic scores can be found in *e*Figure 3 and *eTable 4*.

### Blood-saliva correlations

To further assess the generalizability of epigenetic measures between blood and saliva, we correlated epigenetic clocks and scores between matching blood and saliva samples with and without correction for estimated cell type proportions. As illustrated in *Figure 3*, only 6 out of the 14 tested clocks were highly correlated between tissues (r>.7, p_FDR_<.05), namely PCHorvath, SkinBlood, and PCSkinBlood prior to cell type correction and PCHannum, PCPhenoAge, and PCGrimAge after cell type correction. Notably, correlations between epigenetic age acceleration estimates, which are usually investigated as the main outcomes of interest in epigenetic clock studies, were higher than r=.5 only for two clocks, PCSkinBlood and PCGrimAge. For all other clocks, correlations of age acceleration were moderate to small (r=.20–.44, p_FDR_<.05, cell type corrected).

For all epigenetic scores except BMI (Do et al.), cross-tissue correlations increased after cell type correction, with epigenetic CRP, DNAmTL and PC DNAmTL only showing significant correlations after cell type adjustment. Following cell type correction, both CRP scores and the prenatal smoking exposure score were highly correlated between blood and saliva (r>.7, p_FDR_<.05, cell type corrected). Epigenetic BMI, epigenetic-g, DNAmTL, and PC DNAmTL displayed moderate correlations (r=0.40–0.68, p_FDR_<.05, cell type corrected).

**Figure 3.**
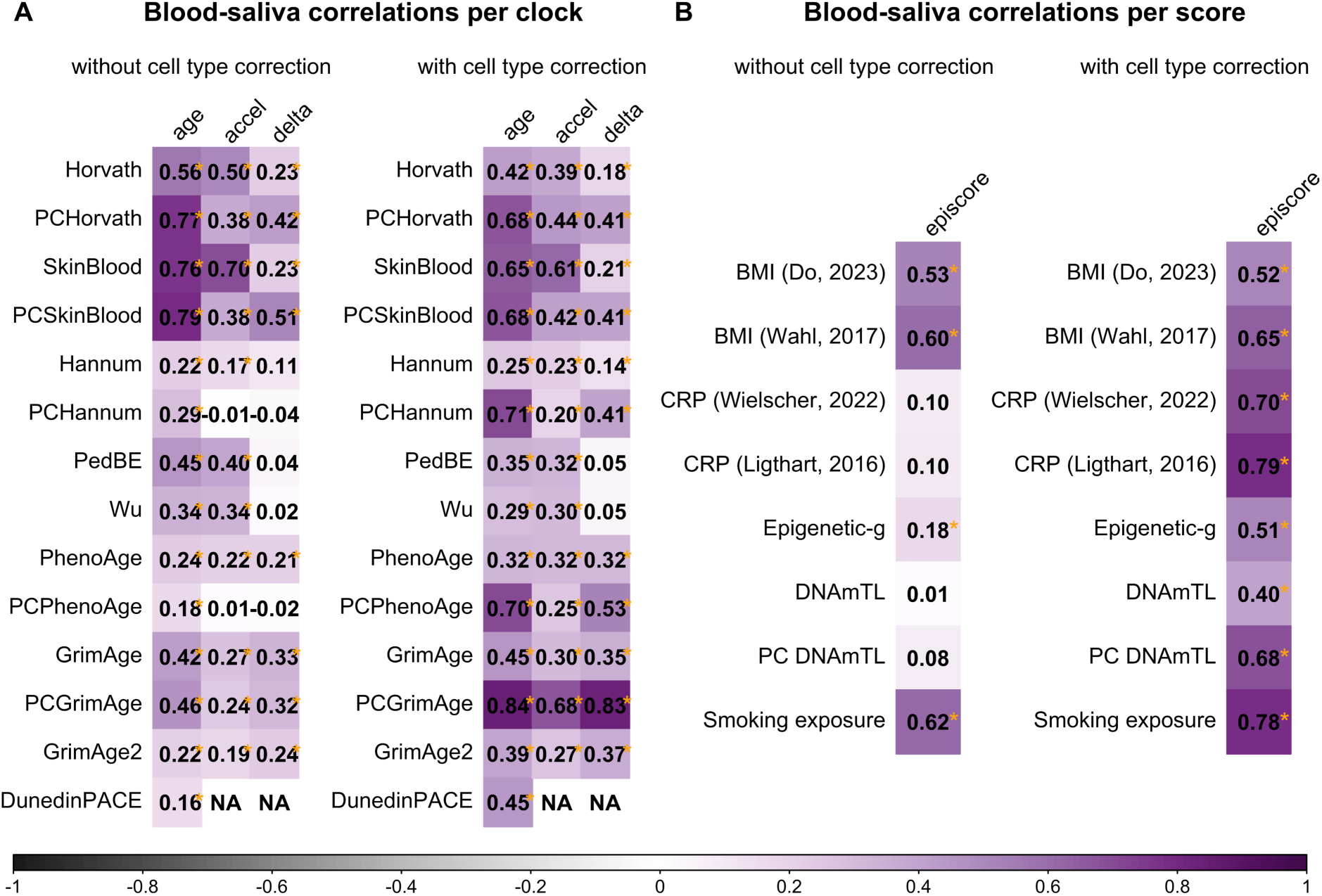
Cross-tissue correlations of epigenetic clocks and scores. Within-person Pearson correlations of epigenetic clocks and scores between blood and saliva. Nominal statistical significance (without multiple testing correction) is denoted by *. Results after FDR correction are available in *eTables 5–6*. All correlations were re-calculated with cell type correction (right side of each panel). *Panel A:* Cross-tissue correlations for epigenetic age (age), epigenetic age residualized against chronological age (accel) and epigenetic age minus chronological age (delta). *Panel B:* Cross-tissue correlations for epigenetic scores.

### Longitudinal stability

Next, we tested longitudinal correlations of clocks and scores between baseline and one-year follow-up within both blood (N=70) and saliva (N=85). Detailed longitudinal results are provided in *eFigure 4* and in *eTables 7–10*. Longitudinal correlations ranged widely (r=.19– .89, cell type corrected) and were higher for epigenetic age than for age acceleration estimates. As expected, PC clocks, which were trained to improve within-person stability, showed higher temporal stability in both tissues (r=.75–.89, cell type corrected) compared with non-PC clocks (r=.19–.61, cell type corrected). Tissue differences were observed for some measures, e.g. PedBE, which showed a higher cross-temporal correlation in saliva than in blood (r_blood_=0.34 vs. r_saliva_=0.69, cell type corrected).

### Effects of early-life adversity and socioeconomic status

Based on previous research showing effects of ELA ^e.g. 60–63^ and socioeconomic disadvantage ^33,34,64,65^ on epigenetic clocks and scores, we wanted to systematically test the generalizability of these associations across pediatric blood and saliva. We detected consistent associations of ELA exposure with higher epigenetic CRP (based on Ligthart, 2016) and lower epigenetic-g, a score trained on cognitive function, surviving FDR correction in both tissues (see *Figure 4*). SES was consistently associated with lower epigenetic-g and accelerated PCPhenoAge, GrimAge, GrimAge2, DunedinPACE, epigenetic BMI (both scores) and epigenetic CRP (both scores) in both blood and saliva. In general, biological epigenetic clocks and epigenetic scores showed higher sensitivity to ELA and SES compared with chronological clocks. For 16 out of 21 tested epigenetic measures, the direction of associations was consistent across tissues. Opposite effect directions in blood and saliva were only observed for non-significant results.

**Figure 4.**
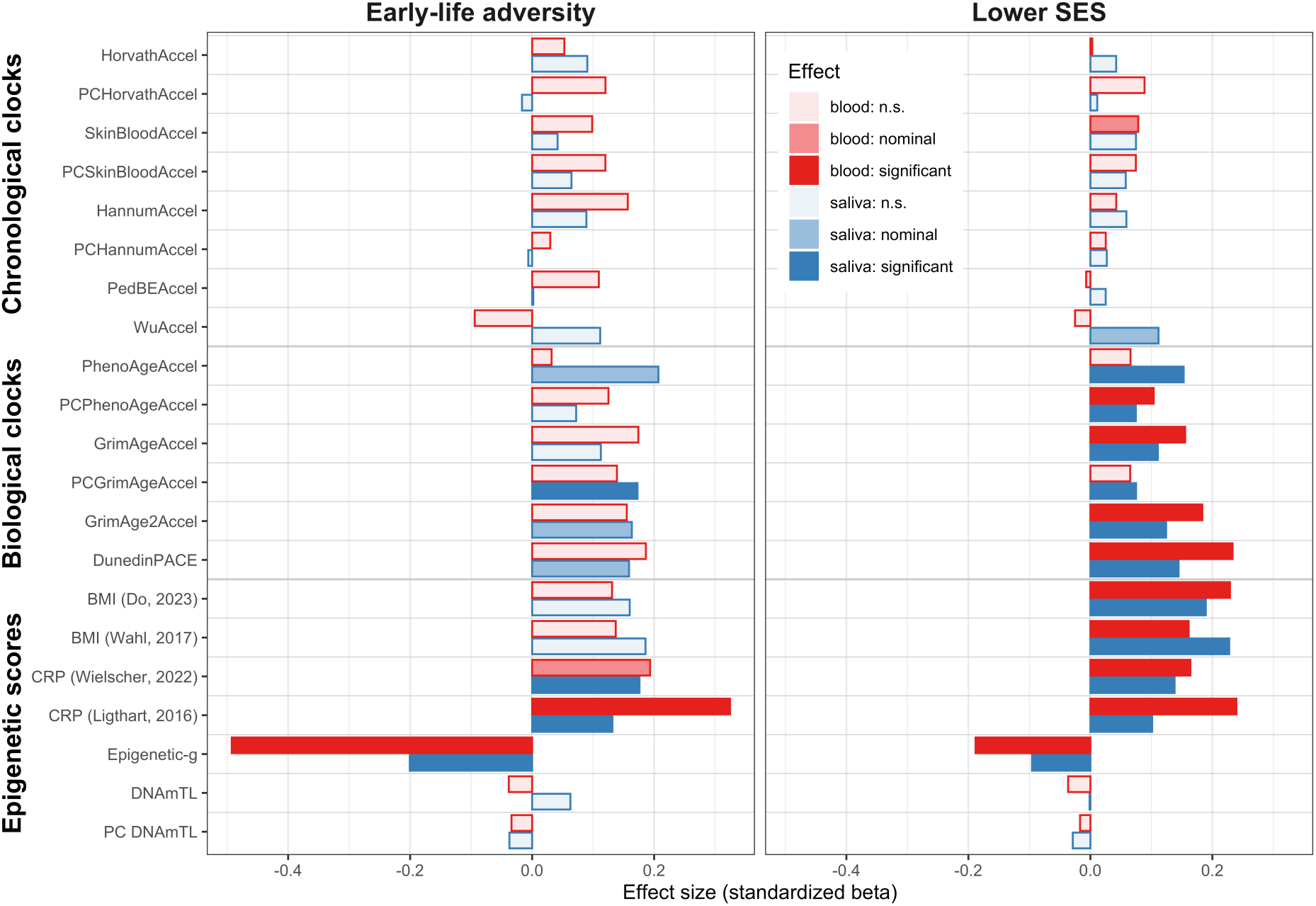
Effects of ELA and SES on epigenetic clocks and scores in blood and saliva. Associations of ELA (early-life adversity; childhood maltreatment or war-/migration-related trauma) and socioeconomic status (SES) with epigenetic age acceleration and epigenetic scores. SES was multiplied with −1 for ease of interpretation. Effects are based on linear regression models corrected for age, sex, genetic kinship, an epigenetic proxy for cigarette smoke exposure (cg05575921), and tissue-specific estimated cell type proportions. Results were FDR-corrected for multiple testing.

### Epigenome-wide analyses

To aid development of valid cross-tissue biomarkers, we investigated blood-saliva correlations for singular CpGs across the entire epigenome. We first calculated within-person Spearman correlations between blood and salivary DNAm for each of 815,069 CpGs available in both tissues. Full results including Spearman correlations and ICC estimates for all CpGs with and without cell type correction are available in *eTable 11*. Only 17.1% of CpGs were significantly correlated between blood and saliva (mean ρ=0.07). After cell type adjustment, this percentage increased to 20.3% (mean ρ=0.08).

In our exploration of the data, we focused on 336,619 CpGs (41.3% of all tested CpG sites) that were variable in both tissues (>5% difference in DNAm between 10^th^ and 90^th^ percentile). The distribution of cross-tissue correlations among variable CpGs is shown in *Figure 5A*: the overall mean correlation was rather low (mean ρ=0.13 without cell type correction, mean ρ=0.15 with cell type correction). We identified a small subset of CpGs (N=19,165, uncorrected; N=27,497, cell type corrected) that were both variable and highly correlated (ρ>0.5) between blood and saliva and made up 2.4% of total the sample without and 3.4% with cell type correction.

Compared with the background of all variable CpGs, highly correlated CpGs were significantly enriched for blood meQTLs ^66^ (OR=5.16, p<2e^−16^ uncorrected, OR=6.45, p<2e^−16^ cell type corrected). CpGs within chronological and biological epigenetic clocks were also significantly enriched for this highly correlated subset (see *Figure 5B*), particularly for biological clock CpGs (OR=3.53, p<.001, cell type corrected). To explore the tissue-specificity of previous epigenome-wide pediatric studies, we extracted mean cross-tissue correlations for trait-associated CpGs from pediatric EWAS ^67^. Correlations were low for all EWAS traits (mean ρ=0.09–0.29, cell type corrected) except for sex (mean ρ=0.39, cell type corrected).

**Figure 5.**
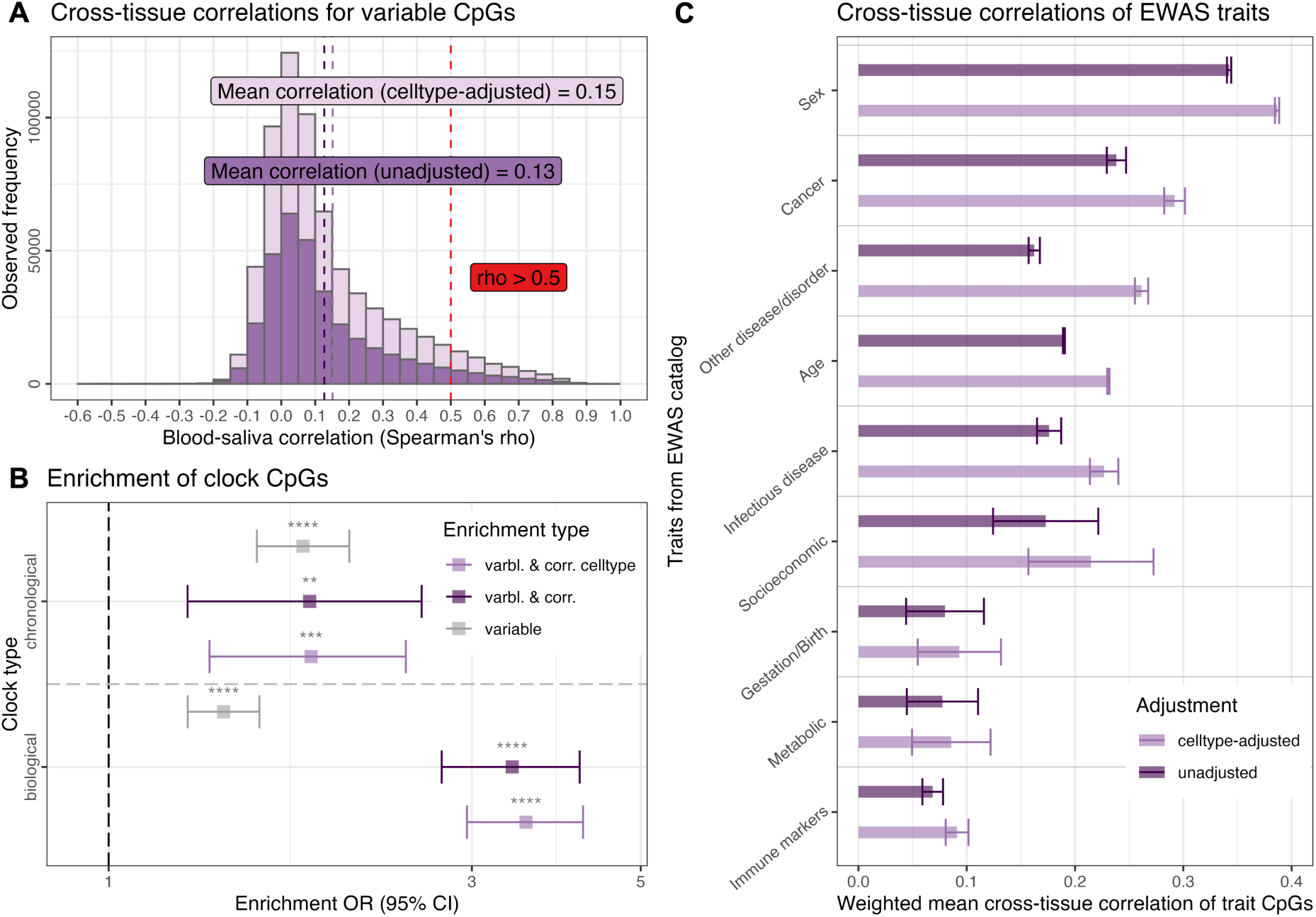
Epigenome-wide cross-tissue analyses. *Panel A:* Distribution of within-person Spearman correlations between blood and saliva for variable CpGs with (mean ρ=0.15) and without cell type adjustment (mean ρ=0.13). Epigenome-wide cross-tissue correlations for individual CpGs are provided in *eTable 11*. *Panel B*: Enrichment odds ratios and 95% confidence intervals of CpGs on chronological (Horvath multi-tissue, Horvath SkinBlood, Hannum, PedBE, Wu) and biological epigenetic clocks (PhenoAge, GrimAge, GrimAge2, DunedinPACE) for the identified subset of variable and highly correlated (ρ>0.5) CpGs. Significance testing is based on Fisher’s exact test. *Panel C*: Mean blood-saliva correlation (± standard error of the weighted mean) for traits from EWAS catalog, based on pediatric EWAS in blood or saliva.

## Discussion

In this study, we evaluated the generalizability of epigenetic measures across pediatric tissues and age ranges. We investigated four questions of relevance for pediatric epigenetics which we will now aim to answer.

### (1) Do established epigenetic measures show sufficient performance in pediatric blood and saliva?

Based on our findings, epigenetic clocks showed moderate to high correlations with chronological age when applied in their training tissue (r=0.3–0.85, cell type corrected), even if they were only trained in older populations. This transfer across ages seems less successful for epigenetic scores. Within the training tissue, correlations with corresponding phenotypes were mostly significant, but low (r=0.06–0.28, cell type corrected). Outside of their training tissue, nine out of 20 evaluated epigenetic measures (Horvath, SkinBlood, Wu’s clock, PhenoAge, GrimAge, GrimAge2, BMI based on Do et al., CRP based on Wielscher et al., epigenetic-g) achieved comparable performance in both blood and saliva after cell type correction.

### (2) How well do epigenetic measures correlate between tissues?

We assessed within-person correlations of epigenetic clocks and scores in matched blood and saliva samples and found that the majority of cross-tissue correlations was moderate to low. Only six out of 22 measures (PCHannum, PCPhenoAge, PCGrimAge, epigenetic CRP based on Ligthart et al. and the prenatal smoking exposure score) score showed high (r≥0.7, cell type corrected) cross-tissue correlations. Concerningly, no epigenetic age acceleration estimate – the most commonly used measure in epigenetic clock studies – achieved a high cross-tissue correlation. PC clocks, which were specifically trained to improve within-person reliability, showed much higher cross-tissue correlations than their non-PC counterparts. Interestingly, cross-tissue correlations improved after cell type adjustment for biological clocks and most epigenetic scores, but worsened for chronological clocks. Our findings are congruent with previous studies ^24,25^, but can now be expanded to a wider pediatric age range.

### (3) Which epigenetic measures are associated with ELA and low SES and is this consistent across tissues?

When investigating associations of ELA and SES with epigenetic age acceleration and epigenetic scores, we observed that the direction of effects was mostly consistent in blood and saliva, even when statistical significance differed between tissues. Associations that survived correction for multiple testing of both ELA and SES were only detected for elevated epigenetic CRP (based on Ligthart et al.^45^) and reduced epigenetic-g, linking to previous findings of elevated inflammation ^68^ and reduced IQ and brain volume ^69^ following childhood maltreatment. These effects were observed in both blood and saliva. Moderate cross-tissue correlation (such as r=0.51 for epigenetic-g) might thus be sufficient to detect consistent exposure effects across tissues. While the clinical relevance of these associations remains to be established, it supports the utility of such measures to identify children at risk, but requires more research and optimization.

### (4) Which CpGs are correlated between blood and saliva across the epigenome?

Optimally, epigenetic clocks and scores for pediatric samples should perform well in blood and saliva. A pre-selection of highly correlated CpGs could support this. We observed that only 17.1% of 815,069 CpGs were significantly correlated between blood and saliva. Among variable CpGs, which are not consistently very highly or lowly methylated across individuals, the average cross-tissue correlation was small (mean ρ=0.15, cell type corrected), similar to previous cross-tissue analyses ^70^. Only 2–3% of CpGs were variable and highly correlated (ρ>0.5) across tissues. This subset was enriched for meQTLs, signifying that DNAm at highly correlated CpGs was also more likely genetically driven. On average, most trait-associated CpGs (based on pediatric EWAS) displayed only low to moderate cross-tissue correlations, indicating tissue-specificity and limited generalizability of EWAS hits.

These insights lead to the final question*: Which epigenetic measures are generalizable in blood and saliva from pediatric populations?* We propose that a well-comparable epigenetic measure should *a)* display a performance pattern according to scenario 1 (equal performance in both tissues after cell type correction) and *b)* show a cross-tissue correlation of r>.7 or higher. In our data, only epigenetic CRP (based on Wlelscher, 2022) fulfilled both criteria. On the other hand, a measure meeting neither of the criteria should not be used outside its training tissue. Based on this, we advise against using PedBE in blood and the Hannum clock, DNAmTL and PC DNAmTL in saliva. The remaining measures either achieved sufficient cross-tissue correlation or equal performance, but not both. For these clocks and scores, careful consideration is warranted for applications outside the training tissue as measures might not reflect the same biological mechanisms in blood and saliva. In summary, our results clearly indicate that additional development of valid cross-tissue epigenetic measures is needed, especially in pediatric populations.

Our dataset of epigenome-wide blood-saliva correlations is intended as a resource for such research. Significant EWAS hits in pediatric populations can be cross-checked against this data to assess tissue-specificity of results. When constructing new cross-tissue epigenetic measures, training data can be filtered based on our dataset to only include features with sufficient cross-tissue comparability – an approach recently applied for training an epigenetic marker of inflammation-related aging ^71^.

This study has a number of limitations. First, blood and saliva DNAm data were generated and pre-processed separately. While within-tissue batch effects were removed, the possibility of confounding due to between-tissue batch effects remains. Moreover, chronological age was chosen as the corresponding phenotype to test the performance of biological clocks, which were not directly trained on chronological age, but on aging- and morbidity-associated biomarkers, biasing our evaluation towards chronological clocks. Additionally, the results of our epigenome-wide analyses are likely to be underpowered and will only capture strong effects. Lastly, our sample is predominantly of European ancestry, meaning that results might not fully generalize to other populations ^72^ and replication in other contexts is warranted.

## Conclusions

Our findings suggest low to moderate generalizability of established epigenetic measures to pediatric blood and saliva, even when cell type composition is taken into account. Future pediatric epigenetic studies should carefully select appropriate tissues and measures. We provide our results as a community resource with the hope of furthering the development of new epigenetic biomarkers in pediatric populations, which may shed new light on the biological pathways between early-life exposures and long-term health outcomes.

## Supporting information

eMethods & eFigures

eTables 1-10

## Funding

This study was funded by the German Federal Ministry of Research, Technology and Space (BMFTR) 01KR1301-A (to CH), 01KR1301-B and 01GL1743-C (to EBB).

## Conflicts of interests

The authors report no conflicts of interest.

## Data availability

Due to the sensitive nature of the data and the informed consent agreements with participants and parents, the Kids2Health data are not publicly available. Interested researchers can obtain a deidentified dataset after approval from the study board. Clock- and score-based analysis results (*eTables 1-10*) are available as *Supplementary Material* and epigenome-wide associations (*eTable 11*) are available on Zenodo: https://doi.org/10.5281/zenodo.17899484

## Code availability

Code for all statistical analyses is available on Zenodo: https://doi.org/10.5281/zenodo.17901642

## Acknowledgements

We would like to thank the children and families who generously participated in this research.

## Notes

### Competing Interest Statement

The authors have declared no competing interest.

### Funding Statement

This study was funded by the German Federal Ministry of Research, Technology and Space (BMFTR) 01KR1301-A (to Christine Heim), 01KR1301-B and 01GL1743-C (to Elisabeth B. Binder).

### Author Declarations

The Ethics Committee of the Ludwig Maximilians University Munich (LMU) gave ethical approval for this work (approval # 18-444).

### Summary of Updates

Corrected typo in 'Statistical Analysis' section; correctly attached & labeled eTables 1-10; correctly labeled eMethods & eFigures; fixed legibility of panel titles in eFigure 3.

