## Supplementary material for "Limited Generalizability of Epigenetic Clocks and Scores Across Pediatric Tissues and Age Ranges": eMethods & eFigures

### eMethods

#### Study population

This study used a subsample of 291 children aged 3-12 years from subprojects one, two, and four of the Kids2Health cohort, a Berlin-based case-control study for studying the mechanisms of biological embedding of early-life adversity. Kids2Health subproject one included 420 children aged 3-12 years, 206 of which had experienced childhood maltreatment as determined by the Maternal Interview for the Classification of Maltreatment (MICM) [1], and 214 age- and sex-matched controls without exposure to maltreatment, violence, or any severe critical or traumatic life events. The MICM was conducted with parents or other primary caregivers and guardians. Severity of abuse and neglect was rated on a 5-point scale. Children were considered maltreatment-exposed based on established cut-off values (emotional abuse $\geq 2$, physical neglect $\geq 2$, physical abuse $\geq 1$, sexual abuse $\geq 1$). Subproject two included 49 children aged 6-12 years with war- and migration-related trauma measured with the UCLA post-traumatic stress disorder reaction index [2]. Subproject four included 66 children aged 3-12 years with clinical obesity (BMI $\geq 95^{th}$ percentile), 39 of which had also experienced childhood maltreatment according to the MICM. Participants were recruited between 2018 and 2022 from local child welfare and protection services, refugee services, pediatric inpatient and outpatient clinics, the Berlin registry of inhabitants, prior studies, and advertisements in schools, newspapers, and online. General exclusion criteria for all children were severe mental or physical disability, severe chronic diseases, chronic intake of medications affecting the nervous system, severe accidents, and insufficient language skills in German or Arabic. Caregivers were compensated monetarily for participation. If required, caregivers received diagnostic results and referral for psychosocial or medical follow-up.

#### DNA methylation

Peripheral blood was sampled between 7:59 am and 10:40 am using PAXgene tubes (QIAGEN) and saliva samples were taken using Oragene DNA collections kits (DNAGenotek) on the same measurement day between 8:45 and 11:34 am, except for three individuals with a maximum difference of 35 days between samples. Blood and salivary DNA were extracted, run on microarrays, and pre-processed separately for each tissue. DNA extraction was conducted in a semi-automated manner using magnetic beads on a Chemagic 360 device (Revvity chemagen Technologie GmbH) using Chemagic DNA Blood Kit H96 and Chemagic DNA Saliva 600 Kit H9. DNA was bisulfite-converted using the EZ-96 DNA Methylation Kit (Zymo Research Corperation) and then run on the EPIC v2.0 Bead Chip (Illumina Inc., San Diego, USA). Assignment of samples to plates and chips was randomized for age, reported sex, ELA status, study subproject, and time point. For children with two time points, both samples were placed on the same chip, but not directly next to each other.

Methylation data were pre-processed in R according to a customized *minfi* (Aryee et al., 2014) workflow (https://github.molgen.mpg.de/mpip/EPIC_Preprocessing_Pipeline). Data were normalized via stratified quantile normalization followed by beta-mixture quantile normalization [3]. We excluded probes on chromosomes X and Y, probes with mapping inaccuracies and flagged probes as reported by Illumina, and probes with a detection p-value > 0.01. For replicate probes, only the probe with the lowest detection p-value was retained. Samples were excluded in case of mean detection p-value > 0.05, mismatches of epigenetically predicted and phenotypic sex, or mismatches of genotypic and methylation data identified with MixupMapper [4]. Blood DNAm data was batch-corrected for plate, chip, and position on chip, and saliva DNAm data was corrected for chip and position on chip using *ComBat* via the *sva* R package [5]. After quality control, 447 samples and 831,713 probes remained in blood, and 501 samples and 825,430 probes in saliva. This resulted in 291 common samples and 815,069 common CpGs across tissues at baseline.

**Epigenetic clocks and scores**

As described in *eTable2*, Horvath, PCHorvath, HorvathSkinBlood, and PCHorvathSkinBlood were originally trained across a wide age range, Wu and PedBE only in children, and Hannum, PC Hannum, PhenoAge, PCPhenoAge, GrimAge2, and DunedinPACE only in (predominantly older) adults. Horvath, HorvathSkinBlood, Hannum, PedBE, Wu, and PhenoAge clocks were calculated using the *methylclock* package (Pelegi-Siso et al., 2021), GrimAge and GrimAge2 with the *meffonym* package[6], and DunedinPACE with the *DunedinPACE* package (Belsky et al., 2022). PC clocks were calculated manually using source code by the authors [7].

Additionally, we computed epigenetic scores by summing the product of CpG weights derived from epigenome-wide association studies (EWAS) for the phenotype of interest with individual DNAm values. The scores were selected based on use in recent studies and sample size of the corresponding EWAS. Epigenetic scores for BMI were calculated using blood EWAS weights by Wahl et al. [8] (used in [9,10]) and a more recent EWAS with a larger sample size [11]. Epigenetic scores for C-reactive protein (CRP) were calculated based on blood EWAS weights by Ligthart et al. [12] (used in [9,13–15]) as well as a more recent EWAS [16]. We calculated an epigenetic score for cognitive ability called *epigenetic-g* (used in [13]). Epigenetically estimated telomere length was calculated based on elastic-net regression-based weights predicting leukocyte telomere length [17] using *methylclock* [18]. An epigenetic score for prenatal smoking exposure was computed using blood-derived EWAS weights [19] (used in [20,21]). Blood cell type deconvolution was performed with a reference panel of twelve immune cell types [22] via *EPIDISH* [23] using the Robust Partial Correlation (RPC) method. Saliva cell type deconvolution was performed in *ewastools* [24] with a reference panel of three salivary cell types specifically designed for children [25]. Blood epigenetic measures were residualized for eleven out of twelve estimated blood cell types (everything but neutrophils, which was the most abundant cell type in the sample) and salivary epigenetic measures were residualized for the estimated proportion of buccal cells.

**Genotyping and relatedness**

DNA was extracted from peripheral whole blood or saliva as described above and genotyping was performed using the Infinium™ Global Screening Array-24 v3.0 BeadChip (Illumina, San Diego, CA). Quality control was performed in PLINK v1.90b7 [26]: single nucleotide polymorphisms (SNPs) with a call rate below 98%, a minor allele frequency below 1%, or deviation from the Hardy-Weinberg equilibrium (p<1e-5) were removed. Individuals with call rates lower than 98% or mismatches between reported and genetically predicted sex were also excluded. MDS (multidimensional scaling) was performed and samples with heterozygosity outliers (> 4 standard deviations from the mean heterozygosity) were removed. 483 samples and 446,540 genetic variants remained after quality control. The sample contained 79 genetically related pairs: three monozygotic twin pairs, 64 full sibling pairs, four half-siblings, and eight cousin pairs, as inferred from identity-by-descent with the PLINK *--genome* command. In order to adjust for the sample's relatedness structure, a genetic kinship matrix was calculated using *PC-Relate* from the *GENESIS* package [27], transformed to the nearest positive definite matrix, and used in sensitivity analyses. All linear regression models containing between-person effects, i.e. all models evaluating clock/score performance, were re-run using the *lmekin* function from the *coxme* package [28] with the genetic kinship matrix as a random effect, excluding two individuals with missing genotype data.

**Salivary and blood biomarkers**

Levels of CRP were determined from saliva using the Salivary C-Reactive Protein Gen II ELISA Kit (Salimetrics, Carlsbad, USA). Assessment was performed in two waves with intra-assay coefficients of variability (CV) of 8.16% (wave 1) and 8.14% (wave 2) and inter-assay CVs of 3.73% (wave 1) and 4.57% (wave 2). CRP levels were log-transformed prior to analysis. Telomere length (T/S ratio) was determined from blood DNA via qPCR run on three plates, with a CV of 5.45% and an ICC of 0.96.

**Additional measures**

Height and weight were measured on the day of biosampling, and used to calculate BMI (N=290). Nonverbal IQ was measured with the SON-R nonverbal intelligence test [29] (N=287). Cognitive testing took place on a separate measurement day up to 244 days after biosampling (mean difference = 24 days). Self-reported ethnicity was assessed as part of the Child Report Kiddie Schedule for Affective Disorders and Schizophrenia (K-SADS) [30] conducted with parents, caregivers, or guardians using the following categories: White, Black, Asian, Mixed: White-Black, Mixed: Asian-Black, Mixed: White-Asian, and Others. Ethnicity information was not available for 13.75% of the baseline cross-tissue sample.

**Epigenome-wide enrichment analyses**

In our epigenome-wide analyses, we identified a subset of CpGs (N=19,165 without cell type correction, N=27,497 with cell type correction) that were variable and highly correlated ($\rho$>0.5) between blood and saliva. In order to further elucidate the function of these CpGs, we performed a set of enrichment analyses. Prior to enrichment, CpGs were mapped to *hg19* based on the Zhou lab annotation (https://github.com/zhou-lab/InfiniumAnnotationV1/raw/main/Anno/EPICv2/EPICv2.hg19.manifest.tsv.gz). Enrichment for blood meQTLs [31] was tested with Fisher's exact test, with all variable CpGs used as background. We then tested whether CpGs on chronological epigenetic clocks (Horvath multi-tissue, Horvath SkinBlood, Hannum, PedBE, Wu) and biological epigenetic clocks (PhenoAge, GrimAge, GrimAge2, DunedinPACE) available on the EPICv2 array were enriched for our set of highly correlated and variable CpGs with and without cell type correction using Fisher's exact test. To ensure that the enrichment was not only driven by overlap with variable CpGs, we also tested for general enrichment of clock CpGs for variable CpGs without consideration of cross-tissue correlations.

In order to investigate the tissue specificity of previous EWAS, we used our resulting database of epigenome-wide blood-saliva correlations to calculate average cross-tissue correlations for traits from the EWAS catalog [32]. 6402 study-specific traits in the catalog were manually annotated to 35 trait categories based on keywords. The catalog was filtered for overlap with the EPICv2 array and only EWAS performed on pediatric blood or saliva/buccal samples with >100 significant hits (according to the p-value threshold of the respective EWAS) and available beta values were retained, resulting in nine remaining trait categories from 5763 studies. EWAS performed in specific blood or salivary cell types were considered as blood-/saliva based. For each trait category, all trait-related EWAS and their absolute beta values of association were extracted. If a CpG was featured in multiple studies for one trait, its beta values were averaged. Then, a weighted average of blood-saliva Spearman correlations for each trait-associated CpG in our epigenome-wide correlation database (*eTable 11*), weighted by the association beta values from the EWAS catalog, was calculated. This analysis was performed based on blood-saliva correlations with and without cell type correction.

**Note on correlation- and agreement-based approaches**

In this paper, we tested blood-saliva comparability using a correlation-based approach, rather than focusing on absolute agreement, as indicated by intraclass correlations (ICC). As noted by Apsley et al. [33], high relative agreement, i.e. a similar rank order of individuals in both tissues, is the most relevant criterion for research settings. For researchers interested in absolute agreement, we also calculated blood-saliva ICCs between blood and saliva for all tested epigenetic measures and CpGs using the ICC function from the psych package [34] (ICC2/random effects model) and provide ICC estimates and their 95% confidence intervals in the *eTables*.

### eFigures

#### eFigure 1. Median absolute error for epigenetic clocks by tissue.

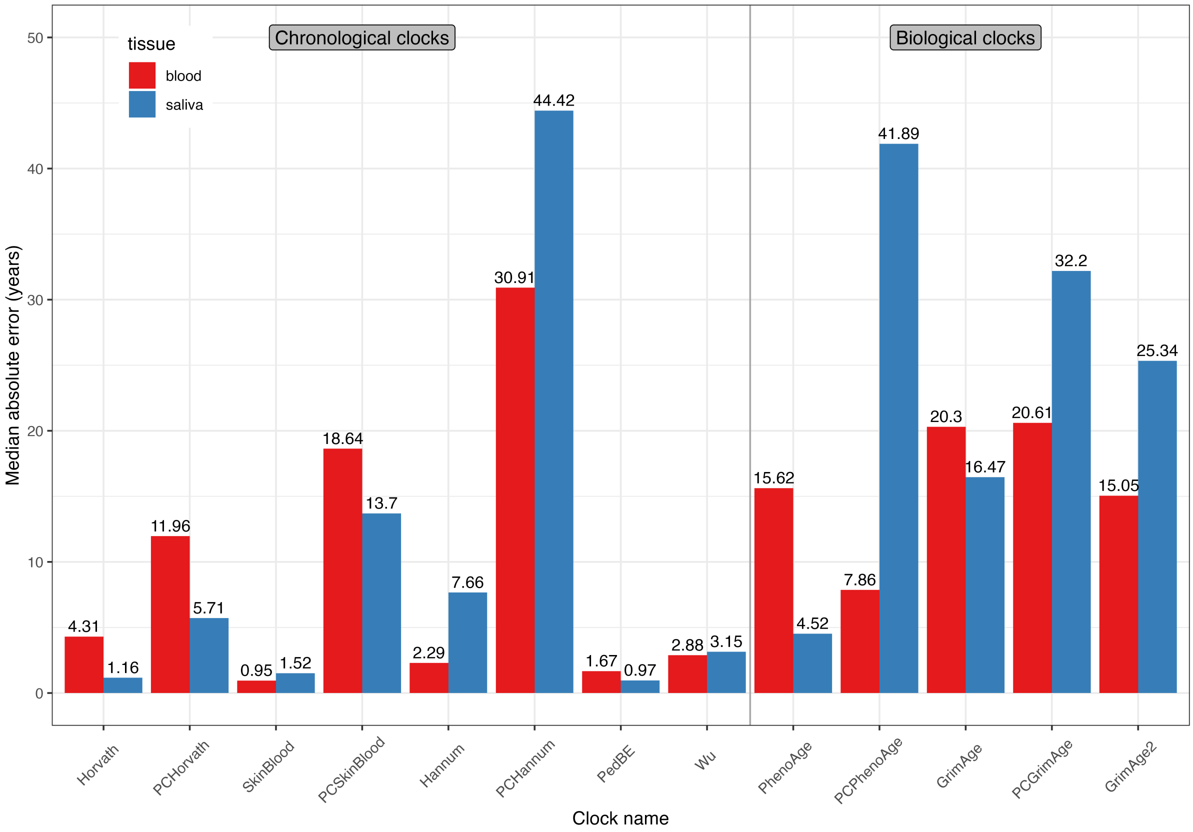

*Legend*. Median absolute error (MAE), reflecting the median of the absolute difference in years between estimated epigenetic and chronological age, by epigenetic clock and tissue.

**eFigure 2. Performance of epigenetic clocks by tissue.**

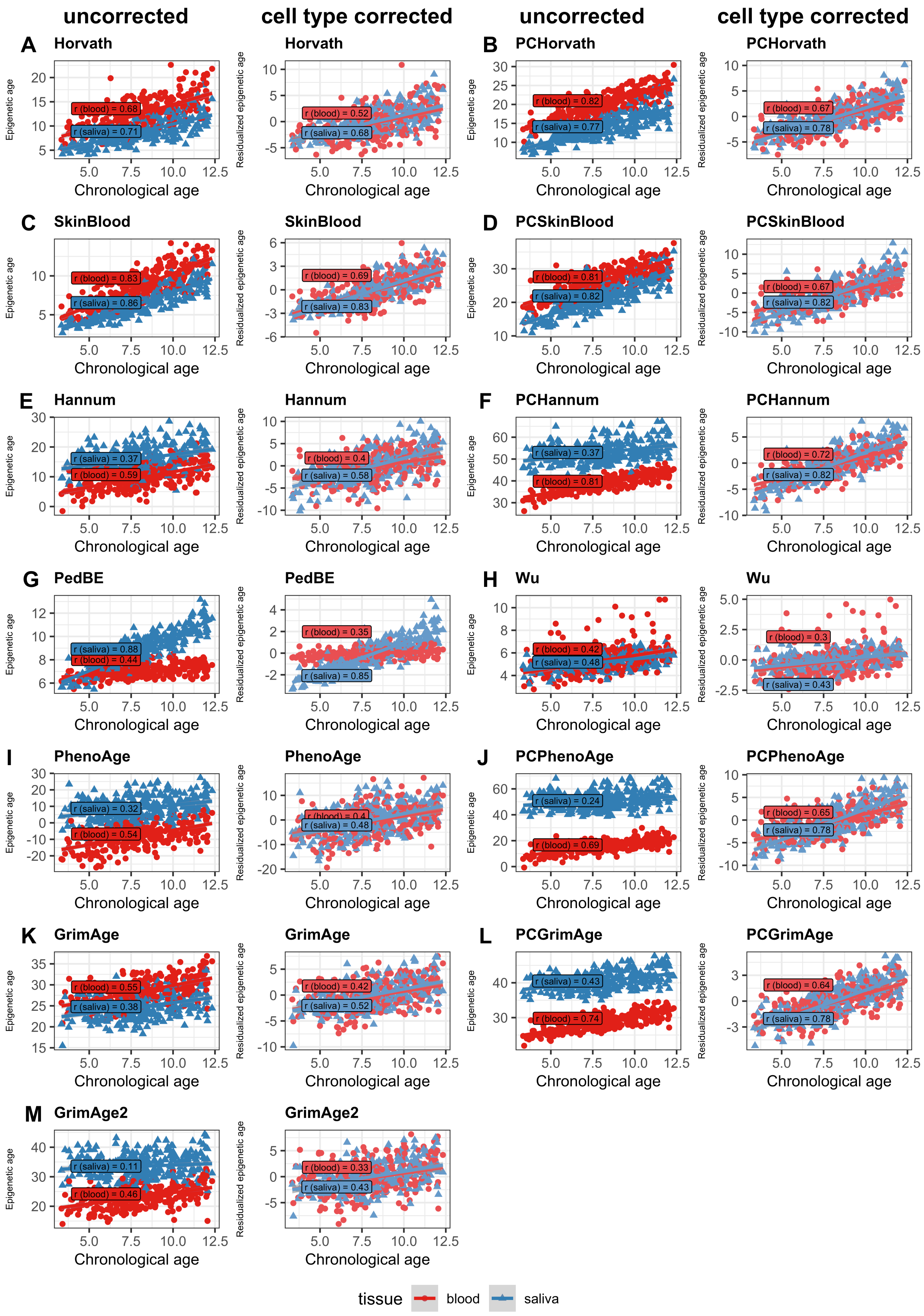

*Legend*. Association between chronological (x-axis) and epigenetic age (y-axis) by tissue for all epigenetic clocks except DunedinPACE. Pearson correlations between epigenetic and chronological age per tissue are labelled as r (blood) and r (saliva). Tissue differences in the intercept indicate a general difference in epigenetic age estimated from blood or saliva, while differences in the slopes indicate differential clock performance between tissues. Results of the corresponding statistical tests of main and interaction effects for each clock are provided in *eTable 3*.

### eFigure 3. Performance of epigenetic scores by tissue.

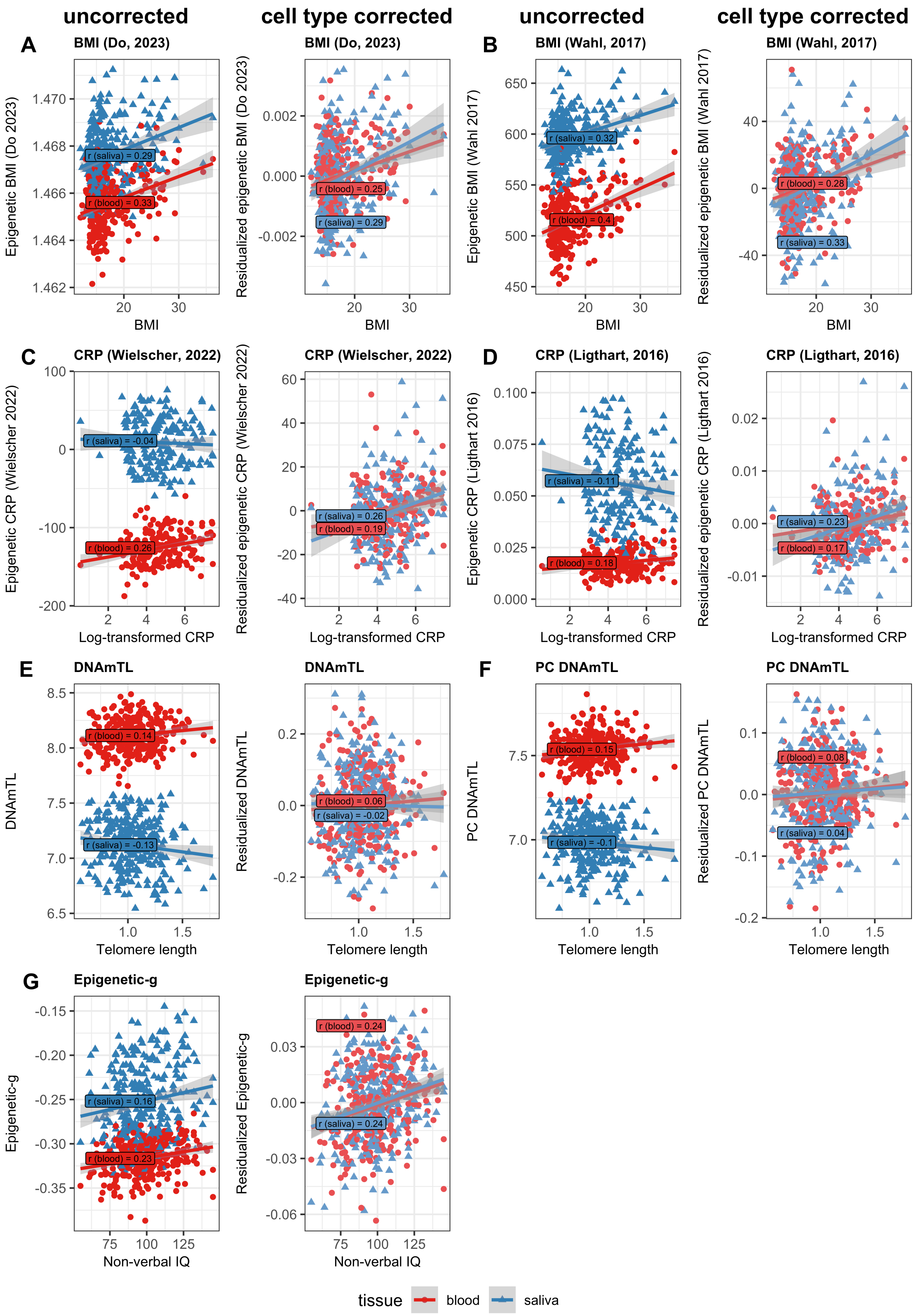

*Legend*. Association between epigenetic scores (y-axis) and the corresponding measured phenotype (x-axis) by tissue for all epigenetic scores with available phenotypes. Pearson correlations between epigenetic score and phenotype per tissue are labelled as r (blood) and r (saliva). Tissue differences in the intercept indicate a general difference in epigenetic scores estimated from blood or saliva, while differences in the slopes indicate differential score performance between tissues. Results of the corresponding statistical tests of main and interaction effects for each score are provided *eTable 4*. CRP was measured in saliva and log-transformed. Telomere length indicates the T/S ratio measured in blood.

### eFigure 4. Longitudinal stability of epigenetic aging measures and epigenetic scores in blood and saliva.

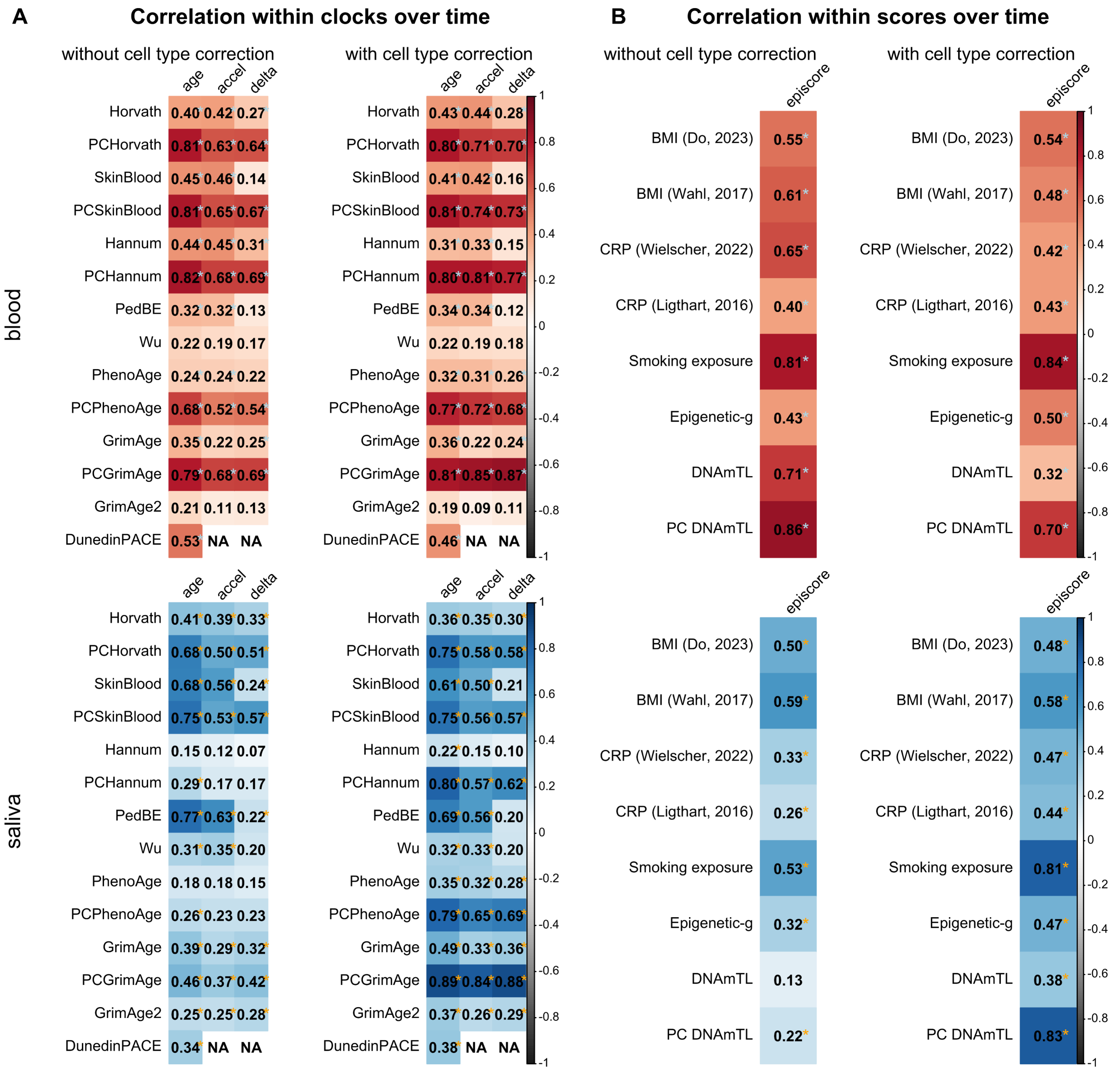

*Legend.* Within-person Spearman correlations of epigenetic aging measures (panel A) and epigenetic scores (panel B) between baseline and one-year follow-up. Correlations were tested separately in blood (N = 70, top, plotted in red) and saliva (N=85, bottom, plotted in blue). Nominally significant correlations are denoted with *. Results after FDR correction and ICC estimates are available in *eTables 7–10*.
